# *Plasmodium* infection induces cross-reactive antibodies to carbohydrate epitopes on the SARS-CoV-2 Spike protein

**DOI:** 10.1101/2021.05.10.21256855

**Authors:** Sarah Lapidus, Feimei Liu, Arnau Casanovas-Massana, Yile Dai, John D. Huck, Carolina Lucas, Jon Klein, Renata B. Filler, Madison S. Strine, Mouhamad Sy, Awa B. Deme, Aida S. Badiane, Baba Dieye, Ibrahima Mbaye Ndiaye, Younous Diedhiou, Amadou Moctar Mbaye, Cheikh Tidiane Diagne, Inés Vigan-Womas, Alassane Mbengue, Bacary D. Sadio, Moussa M. Diagne, Adam J. Moore, Khadidiatou Mangou, Fatoumata Diallo, Seynabou D. Sene, Mariama N. Pouye, Rokhaya Faye, Babacar Diouf, Nivison Nery, Federico Costa, Mitermayer Reis, M. Catherine Muenker, Daniel Z. Hodson, Yannick Mbarga, Ben Z. Katz, Jason R. Andrews, Melissa Campbell, Ariktha Srivathsan, Kathy Kamath, Elisabeth Baum-Jones, Ousmane Faye, Amadou Alpha Sall, Juan Carlos Quintero Vélez, Michael Cappello, Michael Wilson, Choukri Ben-Mamoun, Fabrice A. Somé, Roch K. Dabiré, Carole Else Eboumbou Moukoko, Jean Bosco Ouédraogo, Yap Boum, John Shon, Daouda Ndiaye, Adam Wisnewski, Sunil Parikh, Akiko Iwasaki, Craig B. Wilen, Albert I. Ko, Aaron M. Ring, Amy K. Bei

## Abstract

Individuals with acute malaria infection generated high levels of antibodies that cross-react with the SARS-CoV-2 Spike protein. Cross-reactive antibodies specifically recognized the sialic acid moiety on N-linked glycans of the Spike protein and do not neutralize *in vitro* SARS-CoV-2. Sero-surveillance is critical for monitoring and projecting disease burden and risk during the pandemic; however, routine use of Spike protein-based assays may overestimate SARS-CoV-2 exposure and population-level immunity in malaria-endemic countries.

## Main

Serological surveillance studies provide a fundamental understanding of past exposure to infectious diseases such as SARS-CoV-2. Recent serological surveillance studies reported out of Africa have often reported higher overall seroprevalence than is predicted based on case counts, prompting various hypotheses as to why seropositivity is higher than expected^1–3^.

We identified a high degree of cross-reactivity to SARS-CoV-2 Spike protein (S1 subunit) in individuals with acute (symptomatic and asymptomatic) malaria infection (Figure1A-1B) by ELISA. A total of 741 samples from 617 individuals from 8 countries collected before the first reported case of COVID-19 were tested for Spike protein seropositivity. The observed cross-reactivity was significantly higher in individuals with acute *Plasmodium* infection compared to uninfected individuals in malaria endemic areas (Figure1A-1B). Cross-reactivity was also significantly higher among uninfected individuals living in a malaria endemic setting with previous exposure compared to individuals in a non-endemic settings with no previous malaria exposure.

**Figure 1.**
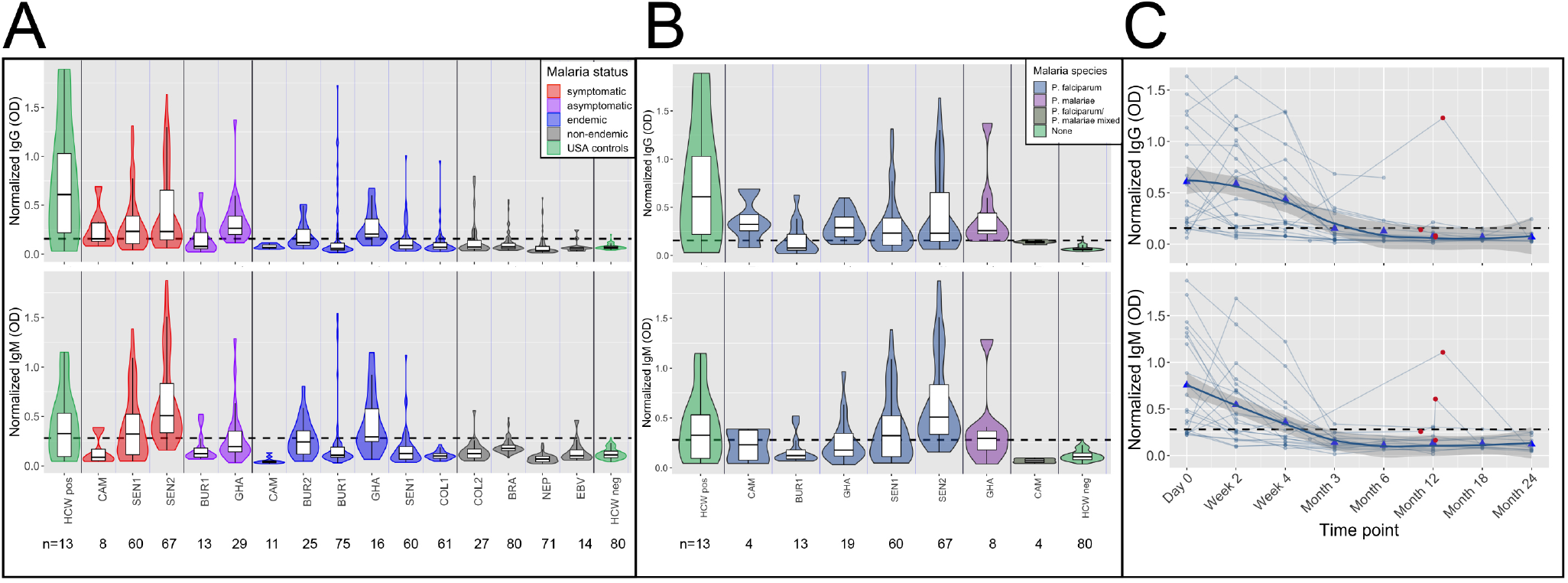
High frequency of cross-reactive antibodies to SARS-CoV-2 Spike protein from *Plasmodium*-infected individuals. In A. and B., Violin plots showing normalized IgG and IgM responses among patients from different cohorts. A. Among all subjects, those in non-malaria endemic areas had significantly lower IgG and IgM than others (t-test IgG p-value<0.0001 and IgM p-value<0.0001.) Subjects with acute malaria infection (symptomatic and asymptomatic) had significantly higher IgG and IgM than uninfected subjects living in malaria endemic areas (t-test p-value<0.0001 for both IgG and IgM). B. Normalized IgG was significantly higher among subjects with *P. falciparum, P. malariae*, and mixed infections than among negative controls (Welch Two Sample t-test p-value<0.0001 for *P. falciparum*, p-value= 0.044 for *P. malariae* and p-value=0.008 for mixed infections), and normalized IgM was significantly higher among subjects with *P. falciparum* and mixed infections but not *P. malariae* than among negative controls (Welch Two Sample t-test p-value<0.0001 for *P. falciparum*, p-value= 0.106 for *P. malariae*, and p-value=0.018 for mixed infections). C. Normalized IgG and IgM over time in 21 subjects with *P. falciparum* monoinfection on Day 0. Both IgG and IgM peaked between Day 0 and Week 4 for all subjects. Reinfection, shown by red circles, boosted IgG response in 1 of 4 subjects and IgM response in 2 of 4 subjects. Bold trend line based on local regression (LOESS). In A. B. and C., normalized IgG or IgM calculated by IgG or IgM OD divided by IgG or IgM of positive control (camelid monoclonal chimeric nanobody VHH72 antibody was IgG control, and pooled convalescent serum from SARS-CoV-2 patients was IgM control). Black dashed lines represent cutoffs for positivity, calculated from normalized IgG and IgM values from 80 RT-qPCR negative HCWs (mean + 3 SDs).

Though patterns of responses were generally similar for IgG and IgM, individuals with symptomatic malaria infection had significantly higher IgM but not IgG than asymptomatic individuals (Welch Two Sample t-test IgG p-value=0.077 and IgM p-value<0.0001). These patterns remain after accounting for age group in a log-transformed multivariate linear regression model. Specifically, children with acute malaria infection had significantly higher normalized IgG and IgM than uninfected children in malaria endemic areas (both p-values<0.0001), and adults with acute infection had significantly higher normalized IgG and IgM than uninfected adults in endemic areas (IgG p-values=0.0047 and IgM p-value=0.0031). In S1-reactive antibody responses measured longitudinally in 131 samples from 21 subjects, IgG and IgM responses peaked between 0-4 weeks post infection for all patients, decreased with time, and were sometimes, though not consistently, boosted by subsequent reinfections (boosting in 1 of 4 IgG samples and 2 of 4 IgM samples with *P. falciparum* (Figure1C).

Of malaria positive subjects, 163 had *Plasmodium falciparum* infection (107 IgG positive and 98 IgM positive), 8 had *P. malariae* infection (6 IgG positive and 4 IgM positive), 6 had *P. falciparum/P. malariae* mixed infections (3 IgG positive and 0 IgM positive), and 1 with *P. vivax* (0 IgG or IgM positive). Normalized IgG was significantly higher among subjects with *P. falciparum, P. malariae*, and mixed infections than among negative controls. Normalized IgM was only significantly higher among subjects with *P. falciparum* and mixed infections than among negative controls. However, the comparison was limited by few subjects with non-*P. falciparum* malaria. Normalized IgG and IgM was not significantly different between subjects with *P. falciparum* and subjects with *P. malariae* (Welch Two-Sample t-test p-value=0.63 for IgG and p-value=0.56 for IgM). Thus, our results suggest that both *P. falciparum* and *P. malariae* induce higher IgG and possibly IgM reactivity.

Since both *P. falciparum* and SARS-CoV-2 infection can induce poly reactive B cells,^4, 5^ we investigated this mechanism as a possible cause of SARS-CoV-2 reactivity. Sera from patients with Epstein Barr Virus (EBV), a disease with characteristic polyreactive B cells responses, were found to have significantly less reactivity than sera with acute malaria infections (t-test p-value<0.0001 for both IgG and IgM for EBV time-points averaging 6 weeks after infection and 6 months after infection), indicating that reactivity is not correlated with poly reactive B cells resulting from EBV infection (Figure1A).

In determining whether cross-reactivity was limited to S1 of SARS-CoV-2 or was observed with other SARS-CoV-2 proteins, we found limited correlated cross-reactivity between Spike S1 IgG and other SARS-CoV-2 proteins (baculovirus expressed Spike ectodomain: Pearson’s R=0.062, p-value= 0.60, and nucleocapsid: Pearson’s R=0.17, p-value=0.15). We also did not find significant correlated cross-reactivity with a commercial test including S2 subunit and nucleocapsid combined (Omega Diagnostics) (Chi-squared with Yates correction p-value= 0.319).

To test whether the cross-reactivity in malaria endemic regions was related to exposure to other alpha and beta-human coronaviruses, we used bacterially expressed peptide arrays^6–8^ and Rapid Exoproteome Antigen Profiling (REAP) for *Saccharomyces cerevisae* expressed exoproteome array of receptor binding domains (RBD) from all seven human coronaviruses, namely SARS-CoV-2, NL63, HKU-1, 229E, SARS-CoV, MERS, and OC43.^9^ Although previous studies suggested cross-reactivity to SARS-CoV-2 is caused by prior exposure to NL63, 229E^10^ and OC43,^11^ we did not observe cross-reactivity between S1 positives by ELISA and peptides, SARS-CoV-2 RBD, or SARS-CoV-2 proteome using either approach except NL63, which showed overall high reactivity (using a REAP Score cutoff of 1.5, 35 of 48 individuals from Senegal and Burkina Faso tested positive [Figure2A]). No samples tested with REAP showed binding to SARS-CoV-2 Spike RBD. Linear peptide epitopes tested against bacterial display libraries did not identify significant binding of malaria-induced antibodies against SARS-CoV-2 peptides. Taken together, these data imply that the cross reactivity could be structural (in the N-terminal domain of S1 not including the RBD) or targeted to a post-translational modification such as carbohydrates.

**Figure 2.**
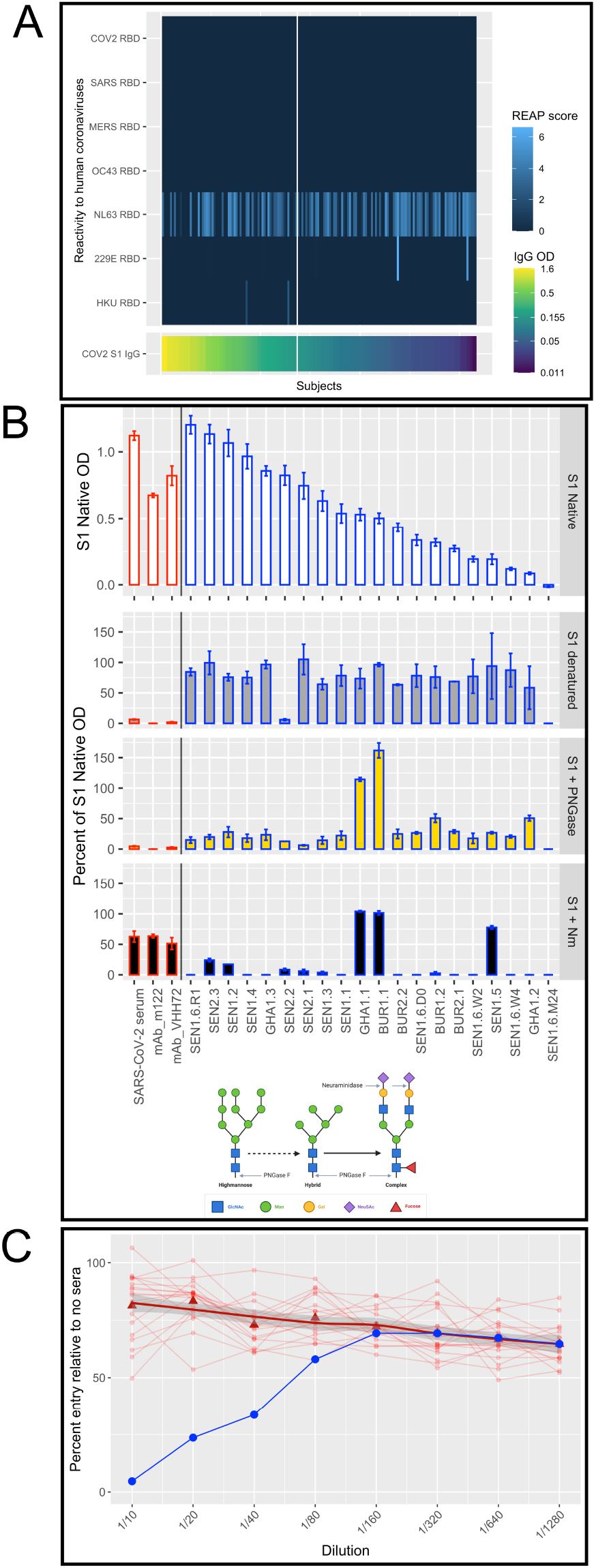
Cross-reactive IgG targets sialic acid on N-linked glycans and is not due to cross-reactivity with other coronaviruses. A. Cross-reactivity to human coronaviruses is only observed for NL63 and differed between S1 positive and S1 negative subjects (S1 positives on the left side of the vertical white line and S1 negatives on the right side) in REAP testing using a REAP score cutoff of 1.5. B. S1 protein was subjected to three conditions to modify the structure: denaturing; treating with PNGase F to remove all N-linked glycans; and treating with Neuraminidase to remove sialic acid. ELISA was performed on each protein condition and the figure shows percent of S1 Native OD under each condition for 3 controls (pooled convalescent serum from SARS-CoV-2 patients, mouse monoclonal M122 antibody, and camelid monoclonal chimeric nanobody VHH72 antibody, all outlined in red) and 20 subjects (outlined in black). Neuraminidase treatment decreased samples to 23.0% (95% CI: 1.1, 44.9) of OD of S1 native, significantly more than SARS-CoV-2 convalescent serum (decreased to 63% of OD of S1 native), suggesting cross-reactivity was due to reactions with terminal sialic from glycosylated sites. C. In a neutralizing assay using pseudotyped viruses (VSV-Renilla luciferase pseudotyped with Spike), 20 samples (red circles) with high S1 Spike IgG ELISA reactivity showed no neutralization. Bold trend line based on local regression (LOESS) of samples. Control (serum from a COVID-19 positive inpatient, in blue) shows neutralization at dilutions of 1/40 and less.

To test the specificity of the cross-reactivity to either structural epitopes or glycosylation, we treated the S1 protein to alter its structure and composition of glycans and carbohydrates. In ELISAs, IgG cross-reactivity was maximally reversed upon treatment with neuraminidase, which removes the terminal sialic acids from glycosylated sites (decreased samples to 23.0% [95% CI: 1.1, 44.9]) S1 native OD, significantly more than SARS-CoV-2 convalescent serum (63% of S1 native OD) (Figure2B). Although treatment with PNGase F (which removes all N-linked glycans) also significantly reduced reactivity (decreased samples to 31.3% [95% CI: 14.3, 48.2] of OD of S1 native), the reduction was less than that of SARS-CoV-2 convalescent serum (decrease to 4.6% of OD of S1 native). Denaturing the S1 protein more modestly reduced reactivity (decreased samples to 71.6% [95% CI: 59.1, 84.1] of OD of S1 native, SARS-CoV-2 convalescent serum decreased to 6.4% of OD of S1 native). These results implicate cross-reacting antibody binding to terminal sialic acids of complex glycans. Of 22-N-linked glycosylation sites spanning the Spike protein, 52% are complex and fucosylated and 15% contain at least one sialic acid.^12^ The majority of the N-linked glycans in the S1 domain of Spike protein (aa16-685) are complex, and are both fucosylated and sialated.^12^ Three samples maintained high reactivity to S1 treated with neuraminidase (133.6%, 126.2%, and 131.2% of OD of S1 native), suggesting that cross-reactivity in these samples could be caused by another mechanism.

We next sought to determine whether these malaria-induced antibodies could neutralize SARS-CoV-2 invasion, possibly providing protection. We performed in vitro neutralization assays testing highly reactive samples using pseudotyped viruses (Vesicular stomatitis virus encoding a Renilla luciferase reporter gene pseudotyped with SARS-CoV-2 Spike) in dilutions from 1:10 to 1:1280 and SARS-CoV-2 viral isolates in Vero-E6 cell plaque assays in dilutions from 1:10 to 1:2430. Similar to a previous study that found no protection from preexisting cross-reactivity,^11^ samples with cross-reactive antibodies did not demonstrate significant neutralizing activity via invasion at any dilution in either experimental system (Figure2C).

Our study reveals that acute malaria infection can cause cross-reactivity to the S1 Spike protein through antibody binding to terminal sialic acids of complex glycans. Reactivity is less pronounced among people previously exposed to malaria but without acute infection. Cross-reactivity was not neutralizing, giving no evidence that malaria exposure protects against SARS-CoV-2 infection through antibody-mediated viral neutralization. Cross-reactivity has also been found in recent studies in malaria-endemic countries; two SARS-CoV-2 serological assays targeting the nucleocapsid protein had cross-reactivity among pre-pandemic samples from Nigeria with higher levels of malaria antibodies^13^. A high rate of false positives was also seen in pre-pandemic samples using commercially available ELISAs from Benin^14^ and from Ghana and Nigeria^15^.

These findings provide evidence of a target and mechanism for cross-reactivity and have implications for the global roll-out of serological surveillance tools that measure IgG or IgM reactive to the Spike protein. Serological surveys remain critical tools in understanding disease burden; however, cross-reactivity in malaria-endemic regions, and especially in patients with acute malaria, could lead to false positive antibody tests for individuals and overestimate population-level exposure. False positive results could cause an individual to underestimate their risk of future infection. At a population level, an overestimate of prior SARS-CoV-2 exposure could lead to overestimates of immunity, meaning larger proportions of people may remain susceptible to infection than serological surveillance might indicate, allowing for continued spread of SARS-CoV-2. Overestimates of exposure would also lead to underestimates of risk of severe disease among people exposed to SARS-CoV-2. Cross-reactive diagnostic tools may have limited utility for serological surveillance when predicting future re-emergence of COVID-19 in areas where incidence of COVID-19 is low relative to malaria. High rates of false positives could preclude using serological tests as correlates of immunity in malaria-endemic areas, hampering policy decision making with respect to risk stratification and implementation of non-pharmaceutical interventions. These results further highlight the need to validate diagnostics in populations with different disease exposures and optimize such diagnostics so that serological surveillance tools can accurately track SARS-CoV-2 exposure.

## Data Availability

Data associated with this manuscript can be found at https://doi.org/10.5061/dryad.w6m905qpj

## Acknowledgements

We would like to sincerely thank the study participants, clinical staff, and field teams for their dedicated involvement in this work. We acknowledge the contributions of Prof. Dyann F. Wirth at the Harvard TH Chan School of Public Health for her longstanding commitment to the collaboration between Harvard and UCAD and her continued mentorship. We are grateful to Elsio A. Wunder, Jr. for technical assistance and coordination for the project.

## Data Accessibility

Data associated with this manuscript can be found at: https://doi.org/10.5061/dryad.w6m905qpj

## Author contributions statement

AKB, AIK, and AMR conceived and directed the project, SL and FL performed the experiments, SL analyzed the data, ACM, YD, JDH, CL, JK, RBF, MSS, MS, ABD, ASB, BD, IMN, YD, AMM, CTD, IVW, AM, BDS, MMD, AJM, KM, FD, SDS, MNP, RF, BD, NN, FC, MR, MCM, DZH, YM, BZK, JRA, MC, AS, KK, EBJ, OF, AAS, JCQV, MC, MW, CBM, FAS, RKD, CEEM, JBO, YB2, JS, DN, AW, SP, AI, CBW, AIK, AMR, AKB performed data acquisition, data analysis and interpretation. AKB and SL wrote the manuscript. All authors have read, provided feedback, and approved the submitted version of the manuscript.

