## Supplemental Figures for "*Plasmodium* infection induces cross-reactive antibodies to carbohydrate epitopes on the SARS-CoV-2 Spike protein"

### This PDF file includes:

Figs. S1 to S2
Table S1

| | | | Malaria exposure status | | | | | | | |
|---|---|---|---|---|---|---|---|---|---|---|
| Cohort | Country | Region | Sympto matic (%) | Asymptomatic (%) | Endemic uninfected (%) | Non-endemic (%) | Male (%) | Mean age, years (min, max) | Dates of collection | # Subjects |
| CAM | Cameroon | Douala | 8 (42.1%) | - | 11 (57.9%) | - | 11 (52.4%) | 26.2 (2, 64) | July-Nov 2018 | 19 |
| SEN1 | Senegal | Kédougou | 60 (50%) | - | 60 (50%) | - | 67 (55.8) | 22 (1, 74) | July 2019 | 120 |
| SEN2 | Senegal | Thiès | 67 (100%) | - | - | - | 67 (100%) | 10.9 (5, 16) | 2015-2017 | 67 |
| BUR1 | Burkina Faso | Bama | - | 13 (14.8%) | 75 (85.2%) | - | 11 (52.4%)* | 2.7 (0.5,4) | July-Aug 2017 | 88 |
| GHA | Ghana | Kintampo | - | 29 (64.4%) | 16 (35.5%) | - | 17 (38.6%)** | 15.1 (3, 70)** | July 2007 and June 2010 | 45 |
| BUR2 | Burkina Faso | Bama | - | - | 25 (100%) | - | 14 (60.9%)*** | 32.6 (21, 43)*** | Oct 2016-Feb 2017 | 25 |
| COL1 | Colombia | Urabá | - | - | 61 (100%) | - | 24 (39.3%) | 31 (5, 80) | Nov 2015-Jan 2016 | 61 |
| COL2 | Colombia | Uramita | - | - | - | 27 (100%) | 6 (22.2%) | 38 (5, 70) | Aug-Sept 2016 | 27 |
| BRA | Brazil | Salvador | - | - | - | 80 (100%) | 30 (37.5%) | 30.6 (5, 71) | Jan-Nov 2010 | 80 |
| NEP | Nepal | Kavrepalanchok and Dolakha | - | - | - | 71 (100%) | 31 (44.3%)**** | 36.2 (4, 80)**** | August 2013-June 2016 | 71 |
| EBV | USA | Illinois | - | - | - | 14 (100%) | 5 (35.7%) | 18.7 (18, 20) | Feb 2015-Oct 2018 | 14 |
| Total | | | 135 (21.9%) | 42 (6.8%) | 248 (40.2%) | 192 (31.1%) | 282 (51.6%) | 22.2 (0.5, 80.6) | July 2007-July 2019 | 617 |

*In BUR1, data on sex was available for 21 of 88 subjects
**GHA had 1 subject with unknown age and sex
*** BUR2 had unknown sex and age for 2 subjects
****NEP had unknown sex and age for 1 subject.

**Table S1. Patient Demographics**

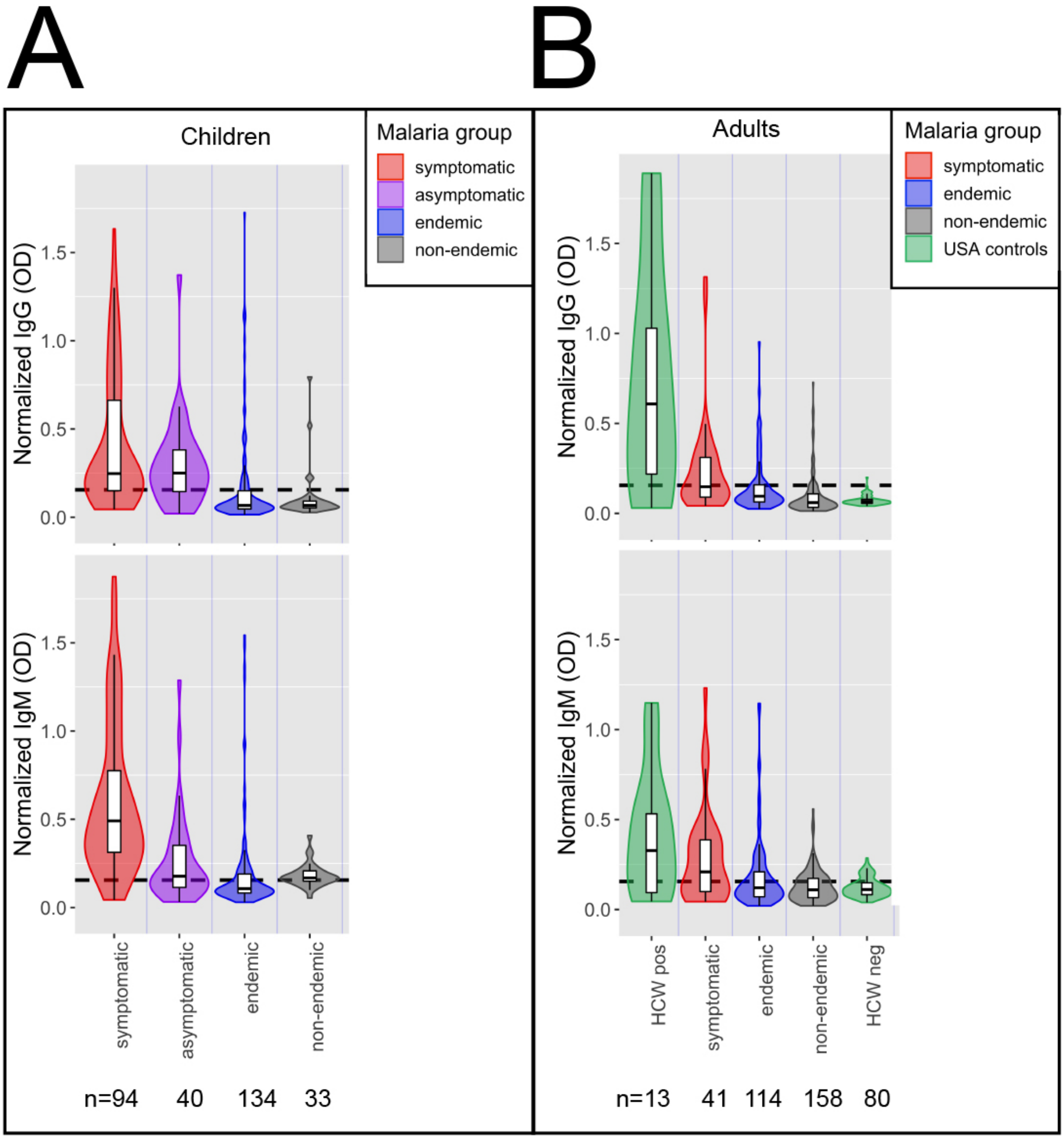


**Fig. S1. Acute malaria infection in both adults and children is associated with S1 subunit Spike cross-reactivity.**
Violin plots showing normalized IgG and IgM responses among a) children and b) adults with different malaria infection statuses. Children with acute malaria infection had significantly higher normalized IgG and IgM than uninfected children in malaria endemic areas (Welch Two Sample t-tests p-values<0.0001 and <0.0001, respectively), and adults with acute infection had significantly higher normalized IgG and IgM than uninfected adults in endemic areas (Welch Two Sample t-tests IgG p-values=0.037 and IgM p-value=0.025). Normalized IgG or IgM calculated by IgG or IgM OD divided by IgG or IgM of positive control (camelid monoclonal chimeric nanobody VHH72 antibody was IgG control, and pooled convalescent serum from SARS-CoV-2 patients was IgM control). Black dashed lines represent cutoffs for positivity, calculated from normalized IgG and IgM values from 80 healthy US healthcare workers controls without SARS-CoV-2 documented exposure (mean + 3 SDs).

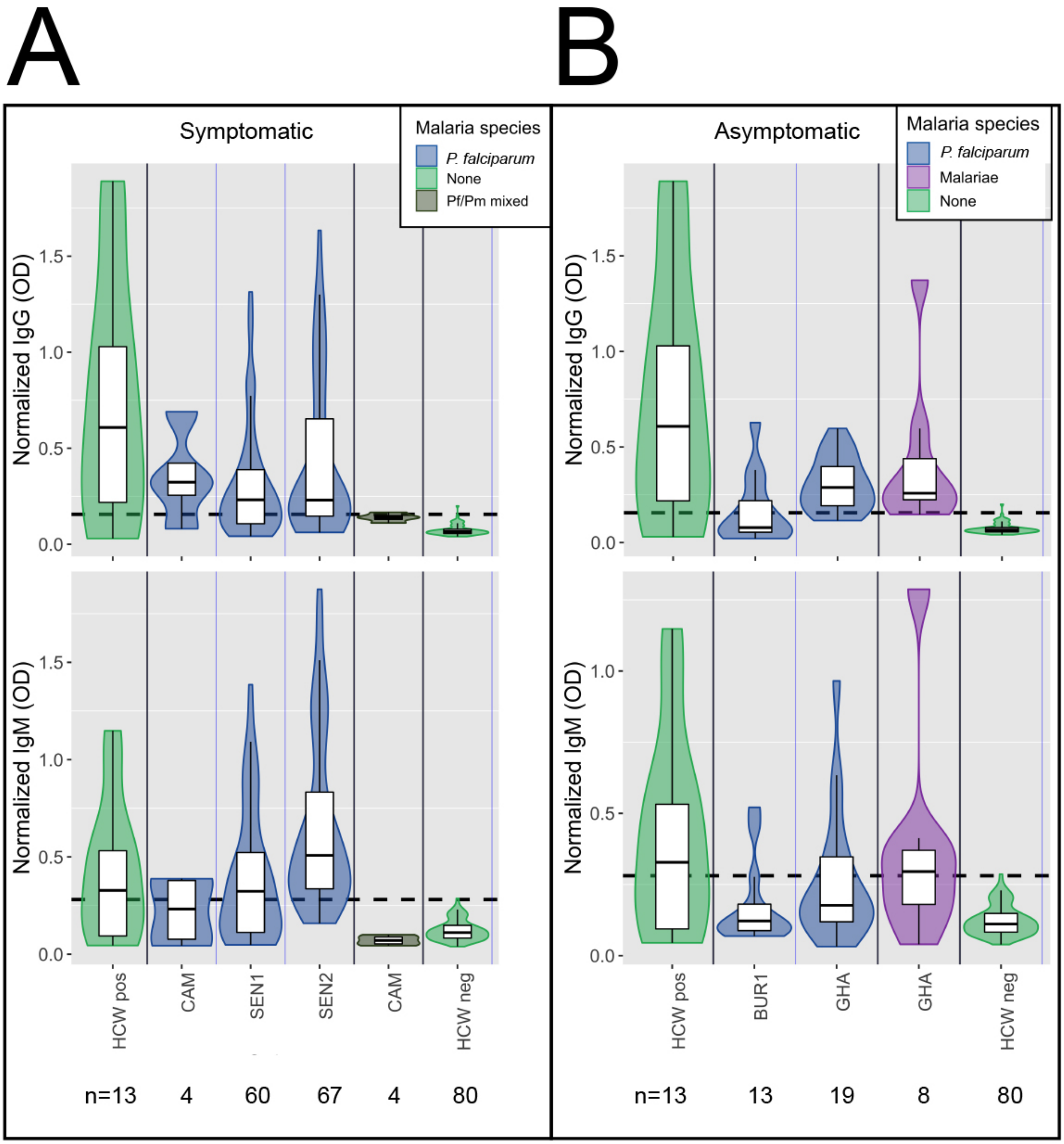


**Fig. S2. Plasmodium falciparum and P. malariae is associated with S1 subunit Spike cross-reactivity in symptomatic and possibly asymptomatic subjects.** Violin plots showing normalized IgG and IgM responses among a) symptomatic and b) asymptomatic subjects by species of malaria infection. For symptomatic patients, both IgG and IgM was significantly higher among subjects with P. falciparum (Welch Two Sample t-tests p-values<0.0001 for both IgG and IgM) and P. falciparum/P. malariae mixed infection (Welch Two Sample t-tests p-values<0.0001 for both IgG and IgM) than healthy US HCWs controls. For asymptomatic patients, both IgG and IgM was significantly higher among subjects with P. falciparum than healthy US HCWs controls (Welch Two Sample t-tests IgG p-value<0.0001 and IgM p-value=0.0063). Asymptomatic patents with P. malariae had significantly higher IgG but not IgM than healthy US HCWs controls (Welch Two Sample t-tests IgG p-values=0.044 and IgM p-value=0.106). Normalized IgG or IgM calculated by IgG or IgM OD divided by IgG or IgM of positive control (camelid monoclonal chimeric nanobody VHH72 antibody was IgG control, and pooled convalescent serum from SARS-CoV-2 patients was IgM control). Black dashed lines represent cutoffs for positivity, calculated from normalized IgG and IgM values from 80 healthy US healthcare workers controls without SARS-CoV-2 documented exposure (mean + 3 SDs).
