## Supplementary material for "*Plasmodium* infection induces cross-reactive antibodies to carbohydrate epitopes on the SARS-CoV-2 Spike protein": Methods

### Ethical statements:

*CAM (Cameroon):* Ethical approval for this study was granted by the University of Doula Institutional Ethics Committee for Research on Human Health (Protocol No 1617), the Institutional Review Board of the Doula Military Hospital (IRB 0180776), and the Human Investigation Committee of Yale University (Protocol 2000023509). All samples were collected with informed consent and in accordance with all ethical requirements of the Institutional Review Boards of the University of Doula, Doula Military Hospital, and Yale University.

*SEN1 (Kédougou, Senegal):* This study was conducted with ethical approval from the National Ethics Committee of Senegal (CNERS) and the Institutional Review Board of the Yale School of Public Health. All research was performed in accordance with relevant guidelines and regulations, and informed consent was obtained from all participants and/or their legal guardians. Samples used in this study were collected as part of ongoing surveillance conducted by Institut Pasteur de Dakar investigating causes of febrile illness.

*SEN2 (Thiès, Senegal):* Ethical approval for this study was granted by the National Ethics Committee of the Ministry of Health in Senegal (Protocol SEN 14/49), the Institutional Review Board of the Harvard T.H. Chan School of Public Health (IRB 14-2830), and the Human Investigation Committee of Yale University (Protocol 2000023287). All samples were collected with informed consent and in accordance with all ethical requirements of the National Ethics Committee of Senegal, Institutional Review Board of the Harvard T.H. Chan School of Public Health, and the Human Investigation Committee of Yale University.

*BUR1 and BUR2 (Burkina Faso):* Ethical approval for these studies was granted by the Institutional Review Boards of Centre Muraz (Ref. A003-2013/CE-CM), the Centre National de la Recherche Scientifique et Technologique (Protocols Ref. A14-2016/CEIRES; A016-2017/CEIRES) and the Human Investigation Committee of Yale University (Protocols 1209010884 and 2000021308). All samples were collected with informed consent and in accordance with all ethical requirements of the Institutional Review Boards of the Centre Muraz, CEIRES, and Yale University.

*GHA (Ghana):* These studies were approved by the Yale University Human Investigations Committee and the Institutional Review Boards at the Noguchi Memorial Institute for Medical Research, the Ghana Health Service, and the Scientific Review Committee and the Institutional Ethics Committee at the Kintampo Health Research Center. In addition, District Ministry of Health representatives and District Ministry of Education representatives approved the study and assisted in communication with participating schools.

*COL1 and COL2 (Colombia):* This study was approved by the Committee of Ethics in Research of the Facultad Nacional de Salud Pública (meeting of 22 May 2014) of the Universidad de Antioquia. All the study participants signed the informed consent.

*BRA (Brazil):* Ethical approval for was granted by the Human Investigation Committee of Yale University (HIC# 1006006956) and from the CONEP in Brazil (CONEP/Brazil 45217415.4.0000.0040).

*NEP (Nepal):* Ethical approval for this study was granted by the Nepal Health Research Council (Reg 106/2013) and the Stanford University Institutional Review Board (IRB 29992). All samples were collected with informed consent and in accordance with all ethical requirements of the Ethical Review Board of the Nepal Health Research Council and the Institutional Review Board of Stanford University.

*EBV (USA):* This study was approved by all relevant the Institutional Review Boards (i.e., those of Northwestern University, the Ann & Robert H Lurie Children's Hospital of Chicago and DePaul University)

*HCW (USA):* This study was approved by the Yale Human Investigation Committee, protocol #2000027690. All samples were collected with informed consent and in accordance with Yale IRB approval.

*YNHH (USA*): A de-identified *P. vivax* infected blood sample was obtained under an approved protocol from the Human Investigation Committee of Yale University.

**Sample collection:**

*CAM:* Samples from Douala, Cameroon were collected from patients aged 6 months or older presenting to the Emergency Department of Douala Military Hospital with fever, history of fever in previous three days, or suspected malaria. Dried blood spots of 150 µL were collected on Whatman™ 3MM chromatography paper, transported at ambient temperature, and then stored at -20°C. Malaria infection and speciation was determined by extracting DNA from dried blood spots (DBS) using Chelex and amplifying DNA with *Plasmodium* genus primers in a Polymerase Chain Reaction (PCR), followed by a second round of PCR with specific species primers of 18S small subunit ribosomal DNA for *P. falciparum*, *vivax*, *malariae*, and *ovale*, as previously described.[1]

*SEN1:* Samples from Kédougou, Senegal, located in Southeastern Senegal 710 km from Dakar, were collected as part of ongoing surveillance conducted by Institut Pasteur de Dakar investigating causes of febrile illness. Patients were recruited from five clinics in Kédougou, Senegal. Eligibility criteria for the main study was the presence of a fever (temperature greater than or equal to 38°C) and/or a fever in the past 24 hours.[2] Positivity for *P. falciparum* was determined based on a *P. falciparum*-specific HRP2/3 rapid diagnostic test (RDT). A venous blood sample of 5ml in a EDTA vacutainer was obtained from consenting, enrolled patients and transported at room temperature from the clinic to the field lab for processing; no more than 6 hours between draw and processing. Thin and thick blood smears were made for each sample to confirm monogenomic infection with *P. falciparum* by microscopy.

*SEN2:* Samples from the low-transmission area of Thiès, Senegal (located about 70 km West of Dakar) come from patients who presented through passive case detection at the Service de Lutte Anti Parasitaire (SLAP) clinic with malaria-like symptoms, tested positive to a malaria rapid diagnostic test (Pfhrp2 antigen RDT), and had positive microscopy for *P. falciparum* monogenomic infection.[3] Participants had a mean parasitemia of 0.77% (range 0.03% to 4.89%).

*BUR1:* Samples from children under five in the high-transmission, rural area of Bama, Burkina Faso were collected during cross-sectional surveys in July and August 2017.[4] Children under 5 provided blood spears and dried blood spots before and after seasonal malaria chemoprevention (SMC). Malaria infection and speciation were determined through PCR from dried blood spots as previously described.[5] Capillary whole blood samples were collected 7-8 days after SMC administration. Blood was centrifuged at 2000xg for 10 min and plasma was collected and stored at -80°C.

*GHA:* Samples from Ghana were collected in 2007 and 2010. In July 2007, four communities in central Ghana were surveyed that were suspected to be endemic for hookworm.[6] The study team randomly selected households to participate, from which up to 2 adults and 3 children were enrolled. Participants submitted approximately 2 mL blood by venipuncture. Two drops of whole blood at the time of collection were used to create thick and thin smears to determine malaria infection and species of infection. The remaining blood was separated by centrifugation and plasma was stored at -80°C.

In June 2010, participants from 16 schools on a 90-km stretch of highway north of Kintampo with height for age (HAZ) of HAZ ≤ −1.80 or HAZ ≥ −0.10 were invited to participate.[7] Blood from participants were tested for malaria with a malaria rapid diagnostic test kit (First Response Malaria Ag HRP-2; Premier Medical Corporation Ltd., Watchung, NJ). Among participants who tested positive via the RDT, species of malaria infection was determined by microscopy. All participants were asymptomatic for malaria at the time of sample collection.

*BUR2:* Participants were enrolled in a cross-sectional study (NIAID R21AI097695) from late October 2016 through February 2017 in the area of Vallée de Kou (Bama) in southwestern Burkina Faso, ~25 km from the city of Bobo-Dioulasso. Malaria is seasonally intense, with the dry season typically beginning in November and lasting through May. Participants were all asymptomatic healthy adults of Mossi or Fulani ethnicity, with negative malaria rapid diagnostic tests and blood smear at the time of sampling, as well as negative HIV rapid testing. Only a single adult from a household was enrolled. Plasma was collected from venous samples, stored in liquid nitrogen after processing, and shipped to Yale by dry shipper in February 2018.

*COL1:* Participants were surveyed in a cross-sectional study on rickettsiae infection from November 2015 to January 2016 in nine hamlets located in Alto de Mulatos in the municipality of Turbo and Las Changas in the municipality of Necoclí in in northwestern Colombia.[8] Residents of all ages were eligible for participation. Households were randomly selected proportional to the number of households in urban and rural areas, and all members of the household were eligible for inclusion. Serum was collected from all participants and stored at -20°C or -80°C. This region is one of the most affected by *P. vivax* malaria in Colombia. *P. falciparum* is also endemic in this region.

*COL2:* Participants were enrolled in a cohort study aimed to study rickettsia infection in Uramita, Colombia from August to September 2016. Participants were selected from households in 10 neighborhoods in the urban center and one village in a rural area. Households were randomly selected proportional to the number of households, and all members of the household were eligible for inclusion. Serum was collected from all participants and stored at -20°C or -80°C. The municipality of Uramita is not considered endemic for malaria transmission.

*BRA:* A prospective cohort among urban slum residents of Pau da Lima, in northwestern Salvador, Brazil was tested for serologic evidence of *Leptospira* infections. This study site is density populated, has low median household per capita income, and a majority of inhabitants do not legally own their domiciles.[9] Households were randomly selected for inclusion. Subjects were eligible for inclusion if they slept at least 3 nights per week in a selected household. Eligible participants provided serum samples in 2010. Malaria is not endemic in this site in Brazil.

*NEP:* This cohort is comprised of subjects who presented to four peri-urban and rural health facilities in Kavrepalanchok and Dolakha, Nepal for a study on enteric fever diagnosis.[10] Subjects had a self-reported >72-hour history of fever, had to be at least 12 months old, and provided a venous blood sample. Although malaria isn't completely eliminated from this area, there is very little malaria transmission in the area and subjects are unlikely to have had malaria.

*EBV:* This cohort contains students who developed infectious mononucleosis caused by Epstein-Barr virus (EBV) during the course of follow-up.[11] A cohort of college students ages 18 to 20 had serum collected at Northwestern University Health Center contributed serum samples from 2 timepoints each within 2 years of diagnosis with EBV. The first timepoint was generally within 6 weeks of the diagnosis of mono (range 6 days before to 5 months after) and the second timepoint was generally 6 months following the diagnosis of mono (range 5 – 24 months).

*HCW pos:* The cohort of health care worker (HCW) positive for SARS-CoV-2 were prospectively followed as part of the Yale Implementing Medical and Public Health Action Against Coronavirus CT (IMPACT) study and tested positive for SARS-CoV-2 during follow-up. This cohort consisted of 13 HCWs who had serum sampled between 15 and 40 days after a positive SARS-CoV-2 diagnosis by RT-qPCR. HCWs tested positive between April 10, 2020 and December 15, 2020.

*HCW neg:* The cohort of health care worker (HCW) negative for SARS-CoV-2 were prospectively followed as part of the Yale Implementing Medical and Public Health Action Against Coronavirus CT (IMPACT) study. This cohort includes 80 HCWs who contributed serum between April 13, 2020 and April 23, 2020 and who had tested negative for SARS-CoV-2 infection by RT-qPCR since the start of the study. Results from this cohort were used to determine cutoffs for S1 subunit IgG and IgM positivity.

*YNHH:* From a patient diagnosed with *P. vivax* treated at Yale New Haven Hospital in Connecticut.

**Experimental methods:**

*S1 subunit ELISA:* Serum (diluted 1:50) was used for all cohorts except CAM, for which dried blood spots were used. For CAM samples, 6mm dried blood spots were eluted in 200μL elution buffer as previously described to obtain the equivalent of 1:40 serum dilution assuming 50% hematocrit.[12] Samples were diluted to a final concentration equivalent to 1:50 serum dilution in dilution buffer (phosphate buffered saline, 0.1% Tween20, and 1% milk powder).

S1 subunit Spike (Acro Biosystems; S1N-C52H2) enzyme-linked immunosorbent assay (ELISA) was performed as described previously,[13] except samples was not treated with Triton X-100 and RNase A, and plates were incubated for 2 hours after adding blocking solution. In addition to samples (diluted 1:50), positive controls for IgG (camelid monoclonal nanobody VHH72-Fc antibody, reactive against SARS-CoV-2 Spike protein,[14] 34 ng/ml, in duplicate), positive controls for IgM (pooled convalescent serum diluted 1:100, in duplicate) and negative controls (serum from healthy patients, from Sigma-Aldrich, in quadruplicate) were included. The optical density (OD) of a sample was then calculated by the value at 450 nm minus the value at 570 nm. Sample OD was normalized by dividing the result by the average of the respective positive control (IgG: camelid monoclonal nanobody VHH72-Fc antibody, 34 ng/ml and IgM: pooled convalescent serum diluted 1:100). Positivity was defined as normalized IgG OD above 0.1557969 and normalized IgM OD above 0.2810049 (mean + 3 standard deviations of the HCW neg cohort, which consisted of 80 healthy US healthcare workers controls without SARS-CoV-2 documented exposure).

*Deglycosylation ELISAs:* ELISAs for S1 treated to different conditions were performed in the same way as the native S1 subunit ELISA, except that the S1 subunit Spike proteins were first treated. S1 native was unaltered. To treat S1 with neuraminidase to remove sialic acid, 2.8μg of native S1 Spike protein was added to 17.2 μl water and 2μl of Neuraminidase stock and incubated at 37°C for 1hr.

For treatment with PNGase F to remove N-linked glycans, 20 μg of S1 native was added to 40 μl $dH_20$. Then, 4 μl 10x Glycoprotein Denaturing Buffer (New England Biosystems) was added, and the mixture was heated to 100°C for 10 minutes. To this mixture, 8μl of 10X GlycoBuffer 2 (New England Biosystems), 8 μl of 10% NP-40 (New England Biosystems), 4 μl PNGase F (New England Biosystems), and 16 μl $H_20$ were added. This mixture was then incubated for 1 hour at 37°C.

For denaturating conditions, 20 μg of S1 native was added to 40 μl $dH_20$. Then, 4 μl 10x Glycoprotein Denaturing Buffer (New England Biosystems) was added, and the mixture was heated to 100°C for 10 minutes.

All proteins treatment conditions were adjusted to a final concentration of 2μg/ml in PBS, the standard coating concentration for the ELISA. Deglycosylation ELISAs were run with three positive controls: pooled convalescent serum diluted 1:100; camelid monoclonal nanobody VHH72-Fc antibody, 34 ng/ml; and Anti-SARS-CoV-2 Spike RBD Antibody, Chimeric mAb, Human IgG1 (S1N-M122) (Acro Biosystems), 8ng/ml. Serum from healthy patients (Sigma-Aldrich) was used as a negative control. Percent of S1 native OD for sample *i* with protein treatment *P* was calculated by: $(OD_{i,P}\text{-}OD_{Neg,P}) / (OD_{i,S1} – OD_{Neg,S1})$ *100%, where OD = IgG OD value, i= sample, P= protein treatment, Neg = negative control, and S1=native S1.

*ELISAs for receptor binding domain of Spike and Nucleocapsid:* 74 samples from the Senegalese cohort (acutely infected timepoints) from Thiès, Senegal (SEN2) were tested by ELISA for ectodomain of Spike and nucleocapsid, as previously described.[15,16] Proteins were expressed in baculovirus cells. Pearson correlations and chi-squared results (using cut-offs for positivity) were found using comparing results between the ELISA for the ectodomain of Spike protein and nucleocapsid protein, and between each of these and S1 subunit Spike protein.

*ELISAs for combined Spike S2 and Nucleocapsid:* 120 samples from the SEN1 cohort from Kédougou, Senegal (acutely infected *Plasmodium falciparum* positive and negative timepoints) were tested with a commercial test including S2 subunit and nucleocapsid combined (Omega Diagnostics Ltd). Chi-squared results (using cut-offs for positivity) were found using comparing results between the combined ELISA and the Spike S1 subunit ELISA.

*REAP:* Rapid Extracellular Antigen Profiling (REAP) was performed as previously described[13] on 131 samples from 21 subjects from Thiès, Senegal (SEN2) and 27 subjects from Burkina Faso (BUR1). This high-throughput discovery method enables detection of antibody reactivities against 2,770 human extracellular proteins displayed on the surface of yeast. Briefly, serum from patients is displayed against this library, IgG coated cells are magnetically isolated, and antigen identity is determined through sequencing. The "REAP Score" is determined from each antibody:antigen binding event, based on how much each protein's barcodes are enriched before and after selection. Samples with REAP Scores of 1.5 and above were considered positive. The receptor binding domain (RBD) of the following coronaviruses were included in the library, as has been done previously: SARS-CoV-1, SARS-CoV-2, MERS-CoV, HCoV-OC43, HCoV-NL63, and HCoV-229E coronaviruses.[13]

*Serimmune:* For identification of antibody binding specificities in 74 samples from 69 subjects from Thiès, Senegal (SEN2), the Serum Epitope Repertoire Analysis (SERA) assay uses a fully random 12-mer peptide library displayed by bacteria as described previously.[17,18] *Escherichia coli* is grown to express a library of $8x10^{10}$ peptides. $8x10^{11}$ *E. coli* cells (10x sampling of the library) are incubated with serum samples on a 96-well, deep-well plate, and antibodies in serum bind to antigen mimic peptides expressed by bacteria.

These results from the SERA assay are then used for the Protein-based Immunome Wide Association Study (PIWAS) to determine associations between immune profiles and exposure to disease, as described previously. Briefly, 12-mer peptides from SERA are divided into 5- and 6-mer (*k*mer) consecutive amino acids, and the enrichment of each *k*mer is calculated by comparing the observed number of reads to the expected number of reads of a particular *k*mer. A PIWAS Score is then calculated for a protein from the enrichment scores of each *k*mer of that protein.

Peptide motifs representing epitopes or mimotopes of malaria-specific antibodies were discovered using the IMUNE algorithm as previously described.[19] IMUNE compared IgG antibody repertoires from subjects with malaria that tested positive on SARS-CoV-2 spike S1 ELISA (n=23) with spike S1 ELISA negative malaria (n=28). Peptide motifs that demonstrated statistical enrichment (Poisson $p<0.01$) in the ELISA positive malaria were then evaluated for cross reactivity to antibody repertoires from SARS-CoV-2 patients from the Serimmune database with serological confirmation of infection (n=378).

*Neutralization with pseudotyped virus (VSV-Spike-RLuc):* 20 samples were selected from the cohort from Thiès, Senegal (SEN2) with a range of S1 Spike IgG values to determine if sera was neutralizing against pseudotyped virus. 293T cells were used to produce Vesicular stomatitis virus (VSV)-based pseudovirus particles. Cells were transfected with a pCAGGS vector encoding Renilla luciferase (Rluc) instead of the VSV G open reading frame. After incubation with inoculum and washing, pseudotyped particles were separated by centrifugation.

$1x10^4$ Vero-E6 cells in 100μl total volume were seeded in black-walled clear bottom 384-well plates (Corning) and incubated overnight at 37°C. The next day, patient sera dilutions were prepared in concentrations of 1:10 to 1:1280 and were pre-incubated with Spike-expressing VSV pseudovirus for 1 hr at room temperature with gentle agitation. Pseudovirus-sera mixtures were added to the adhered Vero-E6 cells at a final virus concentration of 1:10 volume/volume and incubated at 37°C for 24 hrs. Cells were lysed using the Renilla Luciferase Assay System (Promega) according to manufacturer instructions. Luciferase activity was then measured by relative light units using a microplate reader (BioTek Synergy or Cytation 5) and was used to determine percent entry into cells. Percent entry after neutralization was calculated by dividing luminescence at a given serum dilution to that of a no serum control. Means of samples from two runs tested in duplicate were used. For one run, a single replicate of the positive control was successful, thus only one data point is provided in this run. Percent entry among samples was compared against percent entry of a positive neutralization control (serum from a COVID-19 positive inpatient) using a one-sample t-test.

*Neutralization with SARS-CoV-2 virus:* To determine if antibodies in patient serum was protected against wild-type (WT) SARS-CoV-2 virus by neutralization, 10 samples were selected from a weighted group of dynamic range of IgG levels along with negative controls. Neutralization assays were conducted as previously described.[20] Briefly, patient serum was isolated and heat treated. Plasma was diluted from 1:3 to 1:2430 at sixfold serial dilutions and incubated with WT SARS-CoV-2 virus for 1 hr, and then used to infect Vero E6 cells for 1 hr. The serum/virus mixture was removed and cells were incubated and then stained for visualization of plaques at 40 hrs post infection.

*Statistical analysis:* Statistics were performed in RStudio (Version 1.2.5001). Cohorts with 3 or fewer samples were excluded from analysis. Comparisons for continuous variables were performed with Welch Two Sample t-tests and comparisons for binary variables were performed with Chi-squared test with Yate's correction. In all cohorts, subjects were categorized as children if they were <17 years old at their first sampling timepoint. Subjects 17 and older were categorized as adults. For subjects with missing age data, subjects were categorized as adults or children based on the inclusion criteria of the cohort when possible. Statistical tests for age category and malaria status (subjects with acute infection, uninfected subjects in malaria endemic areas, and uninfected subjects in non-endemic areas) were performed in a log-transformed multivariate linear regression model which included interaction terms.
